# Clonal Hematopoiesis of Indeterminate Potential in Crohn’s Disease and Ulcerative Colitis

**DOI:** 10.1101/2024.08.06.24311497

**Authors:** Myvizhi Esai Selvan, Daniel I. Nathan, Daniela Guisado, Giulia Collatuzzo, Sushruta Iruvanti, Paolo Boffetta, John Mascarenhas, Ronald Hoffman, Louis J. Cohen, Bridget K. Marcellino, Zeynep H. Gümüş

## Abstract

Clonal hematopoiesis of indeterminate potential (CHIP) is the presence of somatic mutations in myeloid and lymphoid malignancy genes in the blood cells of individuals without a hematologic malignancy. Inflammation is hypothesized to be a key mediator in the progression of CHIP to hematologic malignancy and patients with CHIP have a high prevalence of inflammatory diseases. This study aimed to identify the prevalence and characteristics of CHIP in patients with inflammatory bowel disease (IBD). We analyzed whole exome sequencing data from 587 Crohn’s disease (CD), 441 ulcerative colitis (UC), and 293 non-IBD controls to assess CHIP prevalence and used logistic regression to study associations with clinical outcomes. Older UC patients (age>45) harbored increased myeloid-CHIP mutations compared to younger patients (age≤45) (*p=*0.01). Lymphoid-CHIP was more prevalent in older IBD patients (*p*=0.007). Young CD patients were found to have myeloid-CHIP with high-risk features. IBD patients with CHIP exhibited unique mutational profiles compared to controls. Steroid use was associated with increased CHIP (*p=*0.05), while anti-TNF therapy was associated with decreased myeloid-CHIP (*p=*0.03). Pathway enrichment analyses indicated overlap between CHIP genes, IBD phenotypes, and inflammatory pathways. Our findings underscore a connection between IBD and CHIP pathophysiology. Patients with IBD and CHIP had unique risk profiles especially among older UC patients and younger CD patients. These findings suggest distinct evolutionary pathways for CHIP in IBD and necessitate awareness among IBD providers and hematologists to identify patients potentially at risk for CHIP-related complications including malignancy, cardiovascular disease and acceleration of their inflammatory disease.

## Introduction

Clonal hematopoiesis of indeterminate potential (CHIP) refers to the presence of age-associated somatic mutations in a proportion of hematopoietic cells of individuals without a hematologic malignancy^1^. CHIP is newly recognized by the World Health Organization Classification of Hematolymphoid Tumors as a distinct entity when detected at a variant allele fraction (VAF) ≥2%^1^. CHIP associated genes are characterized as myeloid-associated genes (M-CHIP) and lymphoid-associated genes (L-CHIP) based on their respective risks for development of myeloid or lymphoid malignancies^2^. While M-CHIP is linked to an increased risk of myeloid malignancies, it is also associated with upregulated inflammatory pathways in mature circulating cells, leading to higher all-cause mortality, chronic kidney disease and cardiovascular disease, and associations with immune diseases including gout and vasculitis^3–9^. Certain features of M-CHIP including specific mutations, clone size as measured by VAF, and rate of expansion are implicated in the risk of progression to malignancy^10^. The detection of CHIP can precede the diagnosis of malignancy by decades^11^.The systemic implications of mutations in lymphoid genes (L-CHIP) are less well understood, but they are similarly associated with an increased risk of lymphoid malignancy^2^.

The pathophysiology of inflammatory bowel disease (IBD) is intimately linked to heightened inflammatory responses and genetic risk variants which are expressed in the lymphoid and myeloid hematopoietic lineages^12–14^. Patients with IBD have an increased incidence of lymphoid and myeloid malignancies which has been associated with exposure to medications and unknown mechanisms^15, 16^. As such, the pathophysiology and clinical course of patients with IBD suggests the need to understand the potential role of CHIP in this population. To date, only a single study has been reported on M-CHIP in IBD and was performed in a modest cohort of ulcerative colitis (UC) patients, identifying a higher prevalence at older ages as compared to historically reported population-based databases^17^. The prevalence of CHIP in Crohn’s disease (CD) has not been reported.

In this study we report new insights into the association between IBD and CHIP, focusing on the overlapping roles of immune cells and inflammatory phenotypes. Towards this end, we utilized the Mount Sinai Crohn’s and Colitis Registry (MSCCR), a richly annotated repository containing detailed clinical and genomic data from individuals with UC, CD, and non-IBD (control) backgrounds. We analyzed deep whole-exome sequencing (WES) data of study participants to identify M-CHIP and L-CHIP mutations as both lineages are implicated in IBD biology. We compared the prevalence of these mutations across different IBD subtypes and non-IBD controls, identifying a high prevalence of M-CHIP in UC and an unexpected presence of CHIP in young IBD patients. Through univariate and multivariate logistic regression, we examined the relationship between CHIP and clinical variables associated with IBD, revealing a strong association between therapy and the risk for CHIP. Furthermore, we highlighted significant gene-pathway associations between CHIP and IBD, suggesting unifying pathobiological mechanisms. Overall, this represents the most extensive investigation of CHIP in the IBD patient population to date and highlights potential clinical implications for IBD patients.

## Methods

### Ethics Declarations

We obtained all necessary institutional review board approvals and oversight was provided by the Program for the Protection of Human Subjects at the Icahn School of Medicine at Mount Sinai. Patients were previously consented under the protocol STUDY-11-01669: “Mount Sinai Crohn’s and Colitis Registry”.

### Sample collection

Participants from the MSSCR, a cross-sectional cohort of 595 CD and 444 UC patients, were recruited between 2013-2016 through visits to outpatient gastroenterology practices, hospital infusion and inpatient visits, and affiliated outpatient endoscopy centers. The 298 control participants in the MSCCR cohort included non-IBD patients visiting the recruitment locations as well as spouses and non-affected family members. Peripheral blood for WES was collected at the time of endoscopy done per study protocol.

### WES processing

WES was performed on the MSSCR cohort of 1,039 IBD patients and 298 controls at the Broad Institute, where the samples were processed as described previously^12, 13^, and CRAM and VCF files were generated.

### WES sample data Quality Control (QC)

To exclude any genetic duplicates, we performed kinship analysis with germline SNPs using KING software and the VCF files^18^. We identified duplicate pairs with kinship coefficient > 0.354 and removed them. In cases where the duplicate pairs were from the same patient, we kept the data from the most recent sample. After sample QC, we included 1,028 IBD patients and 293 controls in our analyses. MSCCR cohort characteristics after sample QC are provided in Table 1.

**Table 1.** Cohort characteristics.

| Phenotype | Category | CD (587)<br># Samples (%) | UC (441)<br># Samples (%) | Controls (293)<br># Samples (%) |
| --- | --- | --- | --- | --- |
| Gender | Male | 331 (56.39%) | 214 (48.5%) | 150 (51.19%) |
|  | Female | 256 (43.61%) | 227 (51.4%) | 143 (48.81%) |
| Smoking status | Smoker | 175 (29.81%) | 141 (31.97%) | 93 (31.74%) |
|  | Never smoker | 412 (70.19%) | 300 (68.03%) | 200 (68.26%) |
| Race | EUR | 496 (84.50%) | 374 (84.81%) | 151 (51.54%) |
|  | AMI | 54 (9.20%) | 33 (7.48%) | 68 (23.21%) |
|  | AFR | 30 (5.11%) | 21 (4.76%) | 65 (22.18%) |
|  | EAS | 7 (1.19%) | 13 (2.95%) | 9 (3.07%) |
| Age (yrs) | <= 40 | 330 (56.22%) | 188 (42.63%) | 35 (11.95%) |
|  | 40 - 50 | 87 (14.82%) | 77 (17.46%) | 38 (12.97%) |
|  | 50 - 60 | 89 (15.16%) | 84 (19.05%) | 154 (52.56%) |
|  | 60 - 70 | 63 (10.73%) | 60 (13.61%) | 57 (19.45%) |
|  | > 70 | 18 (3.07%) | 32 (7.26%) | 9 (3.07%) |

### Somatic Variant Calling

After removing duplicate samples, we called the somatic variants using the Genome Analysis Toolkit (GATK, https://software.broadinstitute.org/gatk) Mutect2 pipeline in tumor-only mode using the preprocessed cram files. Additionally, we provided an external reference of germline variants from gnomAD and a panel of normals (PON) to Mutect2 to exclude likely germline variant calls and sequencing artifacts. To create the PON, we used WES data of 42 young controls (age < 45 years from the control participants) who are unlikely to have CHIP mutations. Next, we applied Mutect2’s orientation bias filter to identify somatic mutations with high confidence and only used those with the PASS filter for further analysis.

### Filtering for Myeloid and Lymphoid CHIP Mutations

Next, to identify M-CHIP and L-CHIP mutation carriers, we filtered for predefined somatic mutations in 76 myeloid and 235 lymphoid driver genes (Supplementary Table S1) at a Mutect2 VAF ≥ 0.02^2, 19, 20^. VAF is defined as the percentage of variant alleles out of the total reads at the sequenced locus. For L-CHIP mutations, in addition to the pathogenic variants in lymphoid driver genes, we also included variants that alter canonical protein sequences as putative markers (Supplementary Table S1)^2^.

### CHIP Quality Control

To filter out false positives, we additionally filtered for mutations with a minimum depth of 20 reads, minimum number of 3 reads supporting variant allele and at least one read in both forward and reverse direction supporting the reference and variant alleles. We also excluded variants observed in gnomAD with allele frequency ≥ 0.001 and variants with observed frequency > 1% in the analyzed cohort unless previously reported to be involved in hematologic malignancies. Next, we annotated the identified somatic mutations for pathogenicity using Combined Annotation Dependent Depletion (CADD) score and excluded mutations with scaled score < 10. For putative L-CHIP mutations, we required additional stringent filtering, which included minimum variant allele read of 5, at least two reads in both forward and reverse direction supporting the variant allele and maximum VAF of 0.2. We reported all individuals with at least one known pathogenic mutation in an M- or L-CHIP driver gene. We provide a complete list of filtered M- and L-CHIP mutations in Supplementary Table S2.

### Statistical Analysis

We first described the prevalence of CHIP mutations in the study population, with focus on distinction by IBD status, type of CHIP mutation and age (Supplementary Table S3). Next, we focused on the VAF of the CHIP mutations by similar stratifications (Supplementary Table S4). Finally, we studied the association between (M- and/or L-) CHIP mutations in IBD patients and demographic and clinical characteristics. A complete list of variables tested is available in Supplementary Table S5. For all association analyses, we computed logistic regression models to calculate the odds ratio (OR) and 95% confidence interval (CI). Univariate logistic regression models were first run to assess possible statistically significant associations. Later, a multivariate logistic regression was fitted with the characteristics identified in the previous step as covariates. All statistical analyses were performed using glm and logistf functions in R.

Based on data availability, we performed stratified analyses within subgroups of CD, UC and non-IBD patients to investigate possible differences in the associations observed. We also analyzed specific IBD phenotype variables, including medical history, surgical history, and medication use across most common categories.

### Data availability

Sequence data used in this study are publicly available in dbGaP Study Accession: phs001642.v1.p1, Center for Common Disease Genomics (CCDG), Autoimmune: Inflammatory Bowel Disease (IBD) Exomes and Genomes. The MSCCR cohort is a sub cohort of participants within this larger study.

### Code availability

The software and computational tools used are described throughout the Methods.

## Results

### CHIP prevalence in IBD

To investigate the overall prevalence of CHIP in IBD patients, we analyzed the peripheral blood WES data from a large cross-sectional cohort of individuals with IBD and controls for their CHIP prevalence. The study design is outlined in Figure 1A, and computational analysis steps for calling CHIP variants are outlined in Figure 1B. In total, we investigated CHIP mutations in 1321 participants which included 587 CD patients, 441 UC patients, and 293 controls. The full demographic information of our study participants is available in Table 1. As the study participants were recruited from the clinic, they were not controlled for age. The median age was 38 years for CD patients, 44 years for UC patients and 54 years for controls. In IBD patients, the overall prevalence of CHIP was 4.57% (47 out of 1,028) (Figure 2A). M-CHIP mutations were identified in 32 IBD patients (3.11%) and L-CHIP mutations in 21 IBD patients (2.04%) patients. In healthy control patients the overall CHIP prevalence was 5.12% (15 out of 293) with M-CHIP mutations in 9 (3.07%) and L-CHIP mutations in 6 (2.05%).

**Figure 1.**
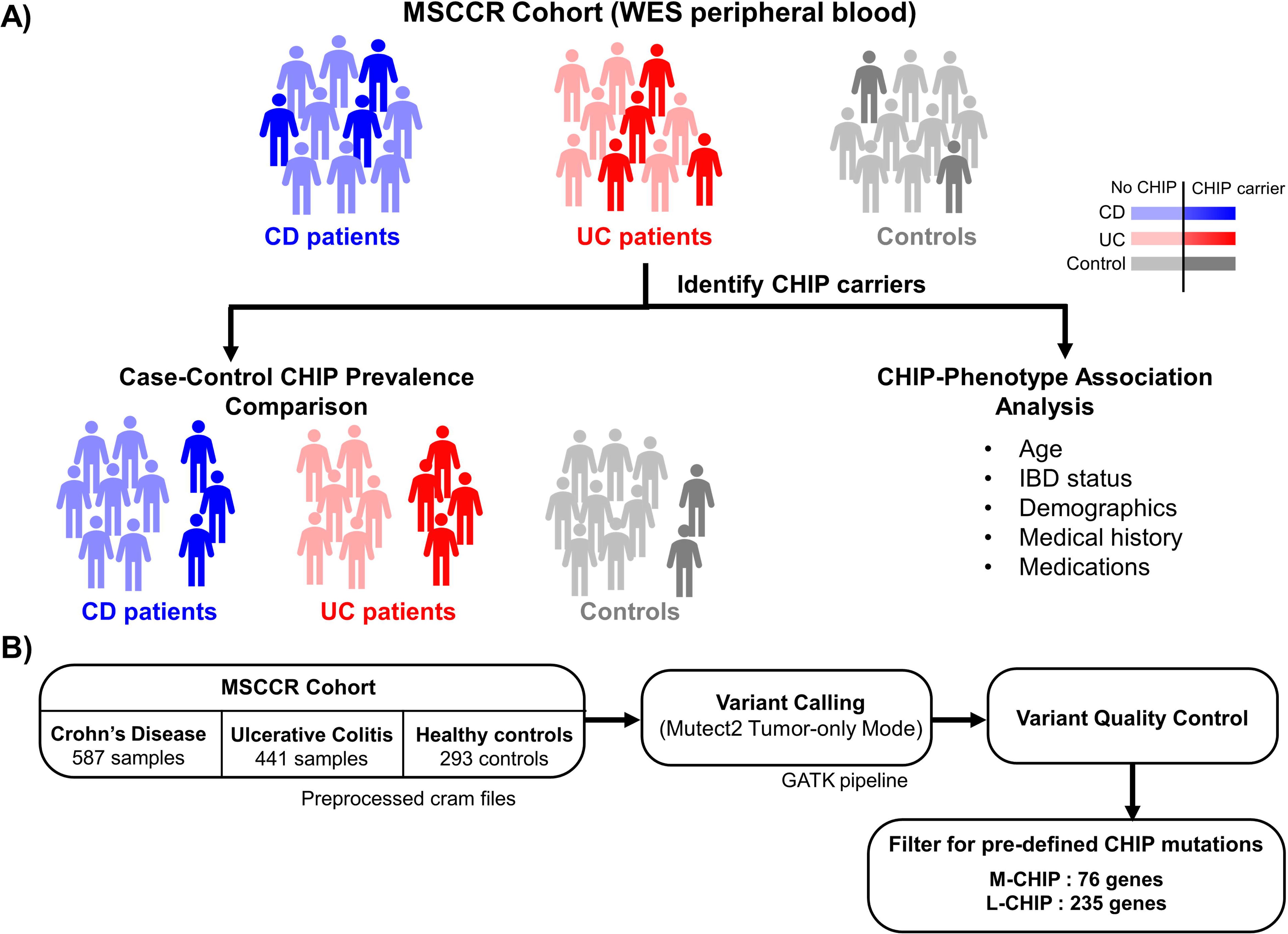
Study design and computational analysis flowchart. A) Study design - We first studied CHIP prevalence in Mount Sinai’s IBD cohort using whole exome sequencing data of IBD patients (CD and UC) and controls. Next, we associated CHIP prevalence with respect to age, demographics and disease characteristics (medications and medical history) B) Pipeline to identify M-CHIP and L-CHIP mutations. Alt text: A diagram with stick figures representing the cohort used in the study and a flow chart describing filtering pipeline for CHIP variant calling.

**Figure 2.**
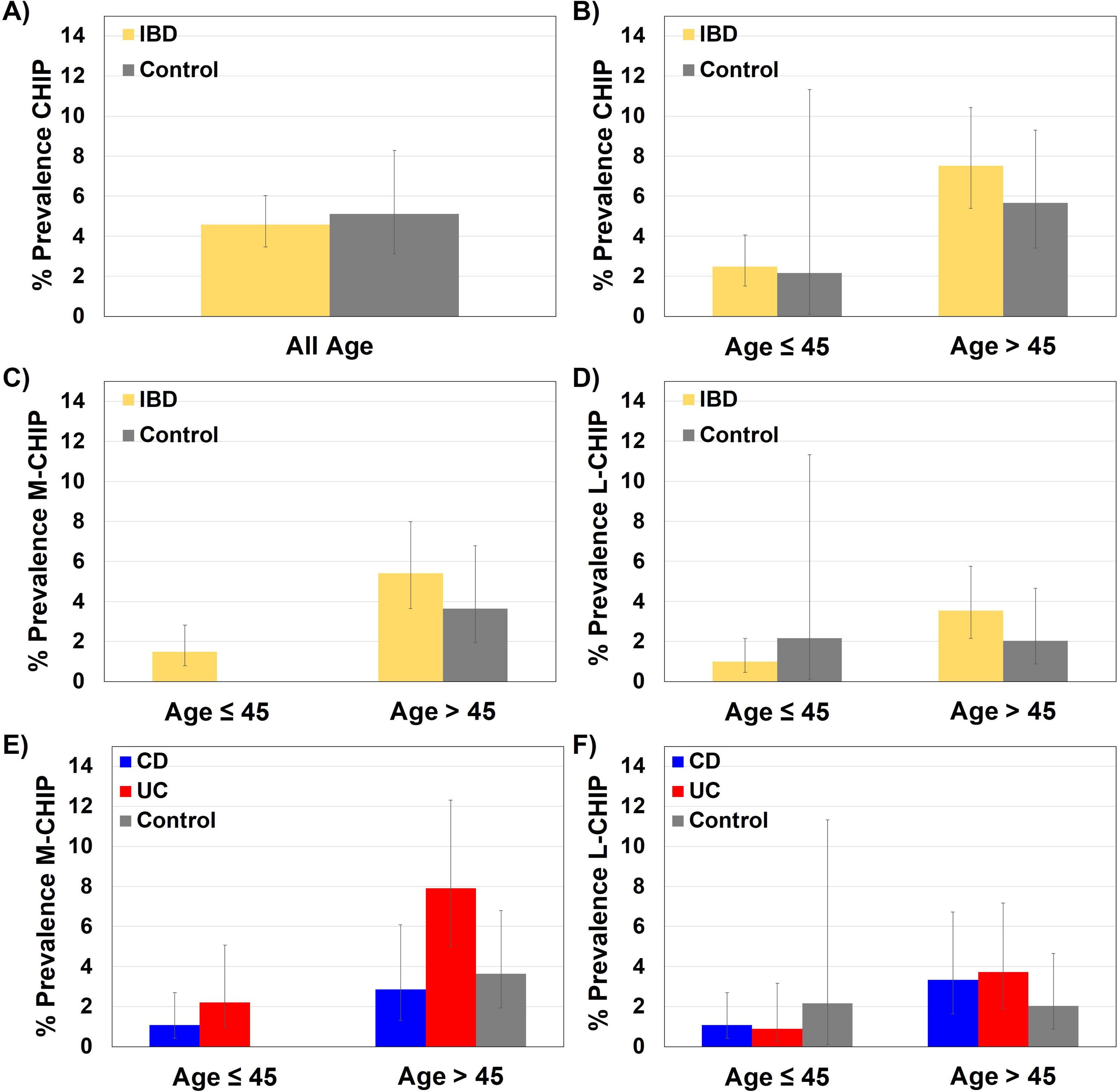
Prevalence of M-CHIP and L-CHIP mutations. Prevalence of CHIP mutations in all IBD patients and controls across all ages (A), and within age cohorts ≤45 and >45 years (B). CHIP was more prevalent in IBD at older ages compared to younger ages (OR 3.19, 95% CI 1.71-5.97, *p* = 2.82E-04). Prevalence of M-CHIP (C) and L-CHIP (D) in all IBD patients and controls for age cohorts ≤45 and >45 years. Both M-CHIP and L-CHIP were more prevalent in older IBD patients compared to younger IBD patients (OR 3.82, 95% CI 1.75-8.35, *p* = 0.001 and OR 3.74, 95% CI 1.44-9.72, *p* = 0.007, respectively). Prevalence of M-CHIP (E) and L-CHIP (F) in CD, UC, and controls by age cohorts ≤45 and >45 years. M-CHIP was more prevalent at older ages in UC when compared to healthy controls (OR 2.27, 95% CI 1.0-5.20, *p* = 0.053) and CD (OR 2.91, 95% CI 1.12-7.53, *p* = 0.028). Alt text: A figure with six panels of bar charts reporting the prevalence of M and L chip in IBD.

### Older UC patients have higher CHIP prevalence

CHIP prevalence is known to strongly correlate with aging^3, 4^. Consistent with this, in older patients with IBD (>45 years old), CHIP was more prevalent in comparison to younger patients (≤45 years old) (OR 3.19, 95% CI 1.71-5.97, *p* = 2.82E-04) (Figure 2B). Most strikingly, among older IBD patients, we observed a higher prevalence of M-CHIP mutations in UC when compared to older healthy controls, and to older patients with CD (OR 2.27, 95% CI 1.0-5.20, *p* = 0.053 and OR 2.91, 95% CI 1.12-7.53, *p* = 0.028 respectively). Both M-CHIP and L-CHIP were more prevalent in older IBD patients (OR 3.82, 95% CI 1.75-8.35, *p* = 0.001 and OR 3.74, 95% CI 1.44-9.72, *p* = 0.007, respectively) (Figures 2C, D). The impact of age on CHIP prevalence in old versus young IBD patients was more pronounced in UC (OR 3.39, 95% CI 1.41-8.14, *p* = 0.006), and less pronounced in CD (OR 2.55, 95% CI 1.01-6.44, *p* = 0.048).

### High-risk CHIP is present in younger patients with IBD

For younger participants (≤45 years), we found that 9 out of 603 patients harbored M-CHIP mutations versus none of the 46 controls (Supplementary Table S3). All CHIP mutations in young patients were M-CHIP mutations, including those commonly identified in the healthy population such as *TET2* and *ASXL1*, suggesting earlier emergence of detectable clones in the younger IBD cohort (Figure 2C).

The clonal burden of CHIP is assessed by VAF, representative of the fraction of CHIP-mutated alleles at a genomic locus identified during sequencing with respect to the overall coverage at that locus. A higher VAF indicates a greater proportion of cells harboring the CHIP mutation, posing an increased risk for CHIP-related adverse events. We assessed whether the clonal burden as measured by VAF of M/L-CHIP mutations was associated with IBD or age. In patients with IBD, younger age (≤45 years) was associated with high VAF for M-CHIP mutations (>0.2 VAF) (OR 8.75, 95% CI 1.47-52.23, *p* = 0.017). In general, CD patients exhibited higher VAF M-CHIP clones compared with UC (OR 6.67, 95% CI 1.18-37.78, *p* = 0.032) (Figure 3A) and younger CD patients had higher VAF M-CHIP mutations compared to older CD patients (OR 33.0, 95% CI 1.90-5507.82, *p* = 0.013) (Figure 3A). There were no IBD or age-specific differences seen with respect to VAF in L-CHIP (Figure 3B, D). This suggests the existence of a subpopulation of young CD with high-risk CHIP mutations that may be at increased risk for hematologic malignancy, and cardiovascular disease and more severe inflammatory outcomes.

**Figure 3.**
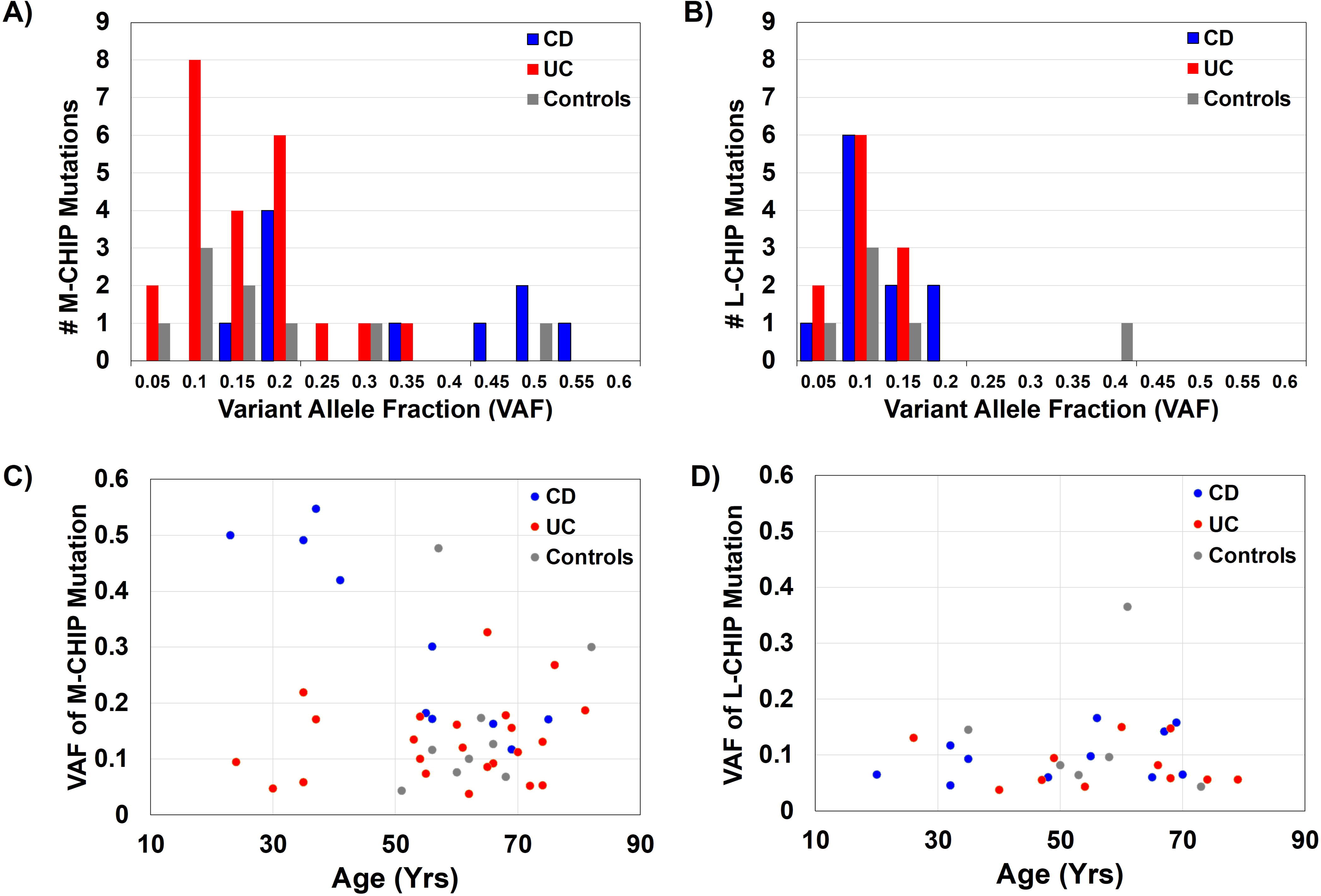
Variant allele fraction (VAF) of M-CHIP and L-CHIP mutations. Histogram of the VAF of M-CHIP mutations (A) and L-CHIP mutations (B). CD patients exhibited higher VAF M-CHIP clones compared with UC (OR 6.67, 95% CI 1.18-37.78, *p* = 0.032). VAF of mutations and age of M-CHIP mutations (C) and L-CHIP mutations (D) observed in UC, CD, and controls. Younger CD patients had higher VAF M-CHIP mutations compared to older CD patients (OR 33.0, 95% CI 1.90-5507.82, *p* = 0.013) Alt text: A four paneled figure reporting variant allele fraction of M and L CHIP mutations and the distribution of VAF across ages of participants.

Unique to those young CD patients was the finding of high VAF *TET2* mutations (Figure 4A, D). We did not observe a difference in the VAF of other common CHIP genes *DNMT3A and ASXL1* across UC, CD, and controls (Figure 4B, C, E, F). The VAF of *TET2* was also markedly higher in CD as compared to UC (Figure 4A), suggesting a relationship between the underlying biology of CD and increased clonal burden of *TET2* CHIP at younger ages. *TET2* encodes for the ten-eleven-transferase-2 protein, which has been previously shown to interact with the glucocorticoid receptor to induce inflammatory signaling in IBD^21^, suggesting that these young CD patients may be subject to more severe inflammatory phenotypes.

**Figure 4.**
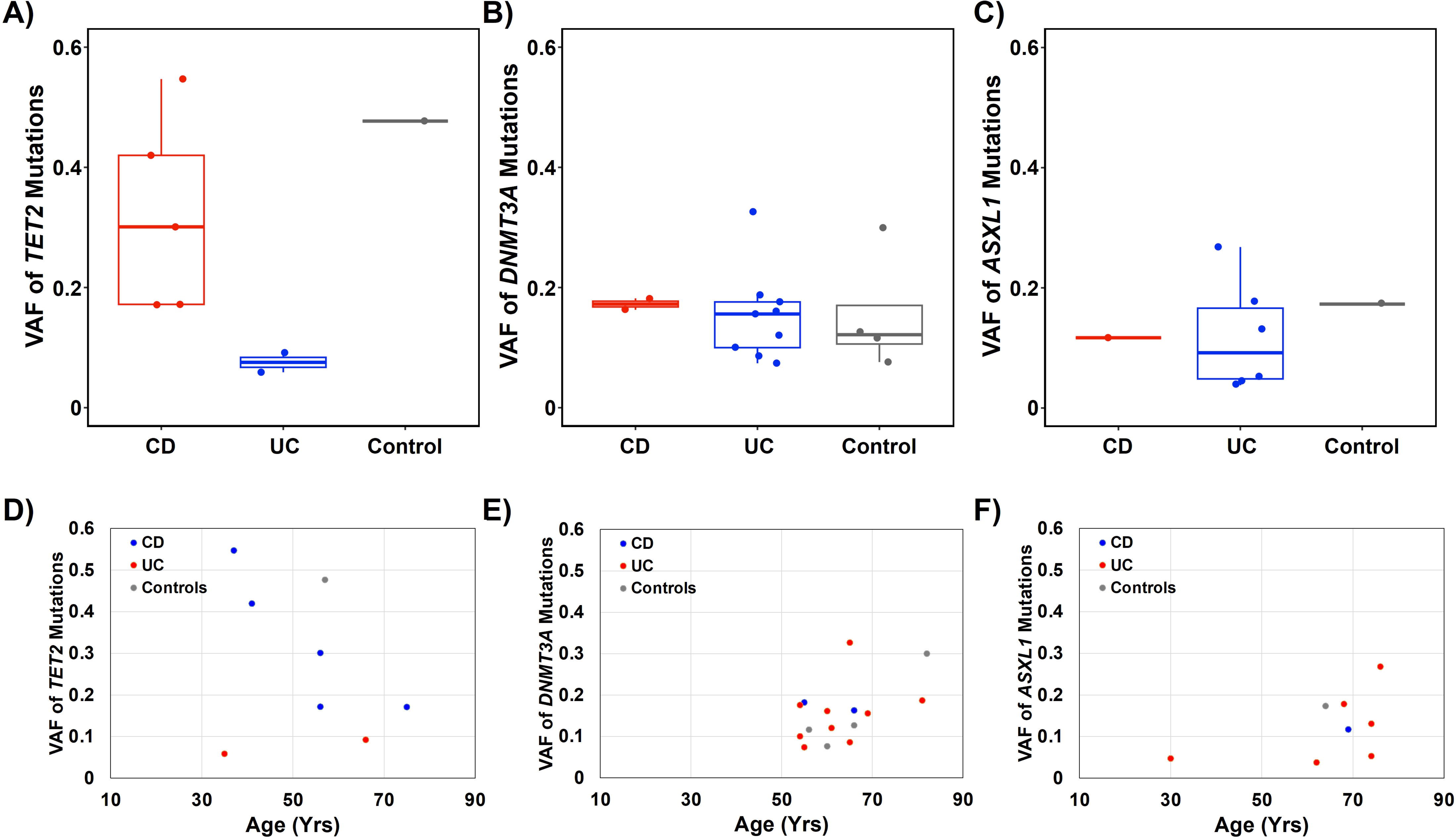
Variant allele fraction (VAF) of mutations in *TET2*, *DNMT3A* and *ASXL1*. Box and whisker plot of VAF for observed mutations within CD, UC, and controls for *TET2* (A), *DNMT3A* (B), and *ASXL1* (C). Comparisons of VAF to age for observed mutations within CD, UC, and controls for *TET2* (D), *DNMT3A* (E), and *ASXL1* (F). Boxplot elements: center line indicates median; box limits represent lower (25th percentile) and upper (75th percentile) quartiles; whiskers extend to 1.5 times the interquartile range. Alt text: A six paneled figure reporting VAF distribution of TET2, ASXL1, and DNMT3A mutations and comparing VAFs across ages for each mutation.

### Patients with IBD have unique CHIP mutations and higher clonal complexity

Analysis of the specific CHIP mutations identified in patients with UC, CD and healthy control patients suggested distinct patterns of gene enrichment across the UC, CD, and control phenotypes (Figure 5, Supplementary Table S2). In UC, of 23 detected M-CHIP variants, *DNMT3A* was the most frequently mutated (9/23), followed by *ASXL1* (6/23) and *PPM1D* (3/23). M-CHIP mutations in *PPM1D* were observed in UC but not in CD, which is consistent with prior reports in UC^17^. In CD, *TET2* was the most prevalent M-CHIP gene (5/10), followed by *DNMT3A* (2/10) (Figure 5B). L-CHIP gene distribution displayed similar segregation between UC, CD, and controls (Figure 5C). *KMT2D* was the most observed mutated gene in UC (3/11), while *CHEK2* and *PCLO* (2/11 each) were the most mutated genes in CD (Figure 5D). Patients with IBD demonstrated greater clonal complexity, defined as the presence of more than one mutation, with 7/47 having multiple CHIP mutations compared to healthy controls of which 0/15 had multiple CHIP mutations.

**Figure 5.**
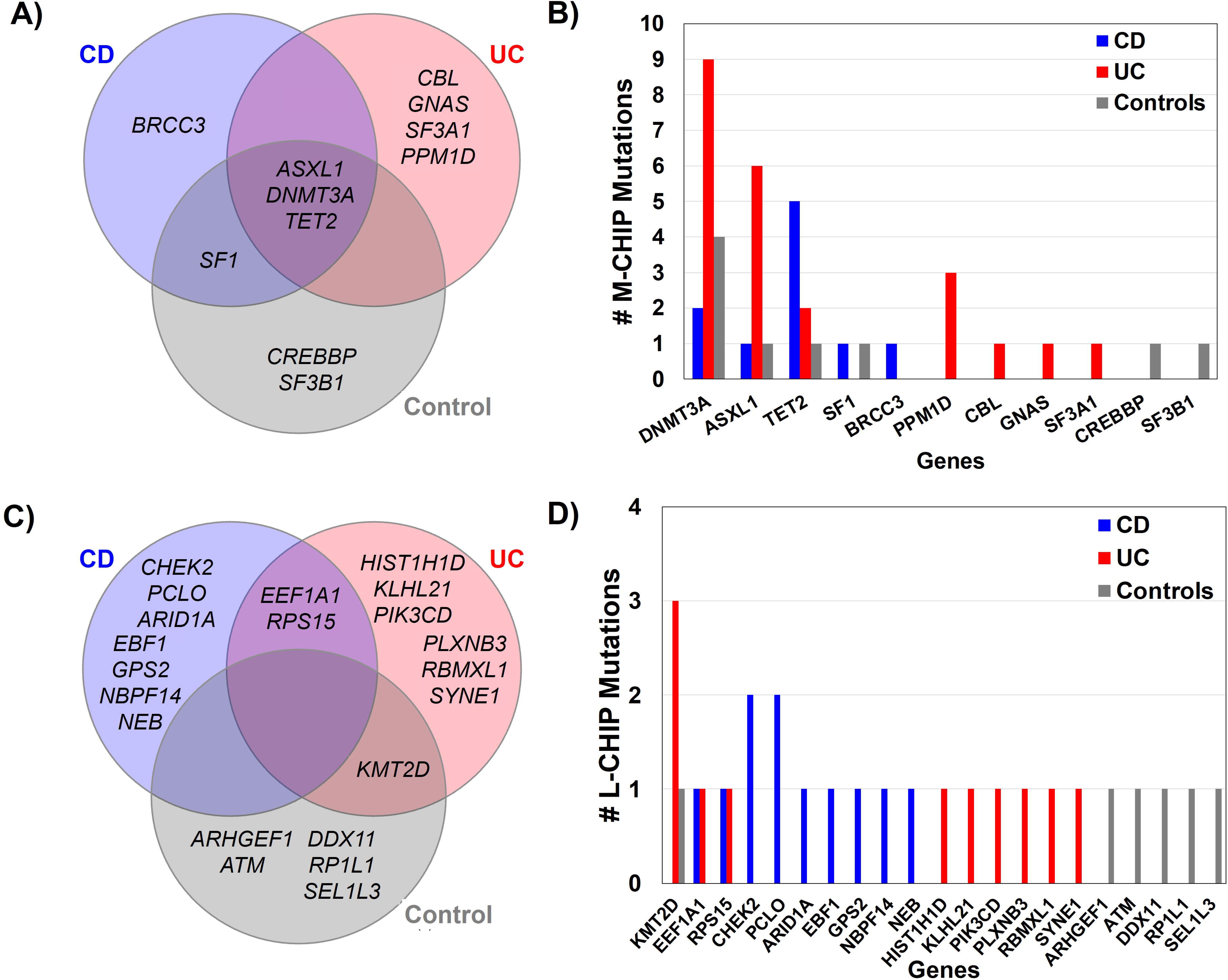
M-CHIP and L-CHIP genes in CD, UC and controls. Venn diagram showing overlap of specific genes identified in CD, UC, and controls for M-CHIP (A) and L-CHIP (C). Number of mutations identified in M-CHIP genes (B) and L-CHIP genes (D) categorized by CD, UC, and controls. Alt text: A four paneled figure including Venn diagrams showing overlap of genes identified in each IBD subtype, and frequency of genes across each subtype.

### Ancestry and IBD therapy were associated with the presence of CHIP

To understand the relationship between CHIP and IBD pathophysiology we assessed the relationship between IBD patient demographics, medication exposures, and disease outcomes with the prevalence of CHIP mutations in a univariate model (full list of tested variables is available in Supplementary Table S5).

Among demographic variables, age was a significant risk factor for CHIP prevalence as previously described. Ancestry was also found to be a risk factor for CHIP. Using European ancestry as a reference variable in our univariate model, participants of East Asian ancestry were at higher risk of CHIP (OR 3.97, 95% CI 1.11-14.16, *p* = 0.03) and participants of African American ancestry were at increased risk of M-CHIP (OR 3.77, 95% CI 1.38-10.34, *p* = 0.01).

Current corticosteroid use was positively associated with the presence of CHIP (OR 2.03, 95% CI 1.00-4.12, *p* = 0.049) and L-CHIP (OR 3.13, 95% CI 1.31-7.48, *p* = 0.01) while current use of anti-TNF therapies was negatively associated with the prevalence of M-CHIP (OR 0.31, 95% CI 0.11-0.89, *p* = 0.03) (Figure 6A). In multivariate analyses taking into account age and ancestry, current use of steroids remained a statistically significant risk factor for CHIP (OR 2.10, 95% CI 1.00-4.40, *p* = 0.049) and for L-CHIP (OR 3.36, 95% CI 1.32-8.54, *p* = 0.01), further drawing attention to the potential link between a treatment refractory IBD phenotype and CHIP mutations.

**Figure 6.**
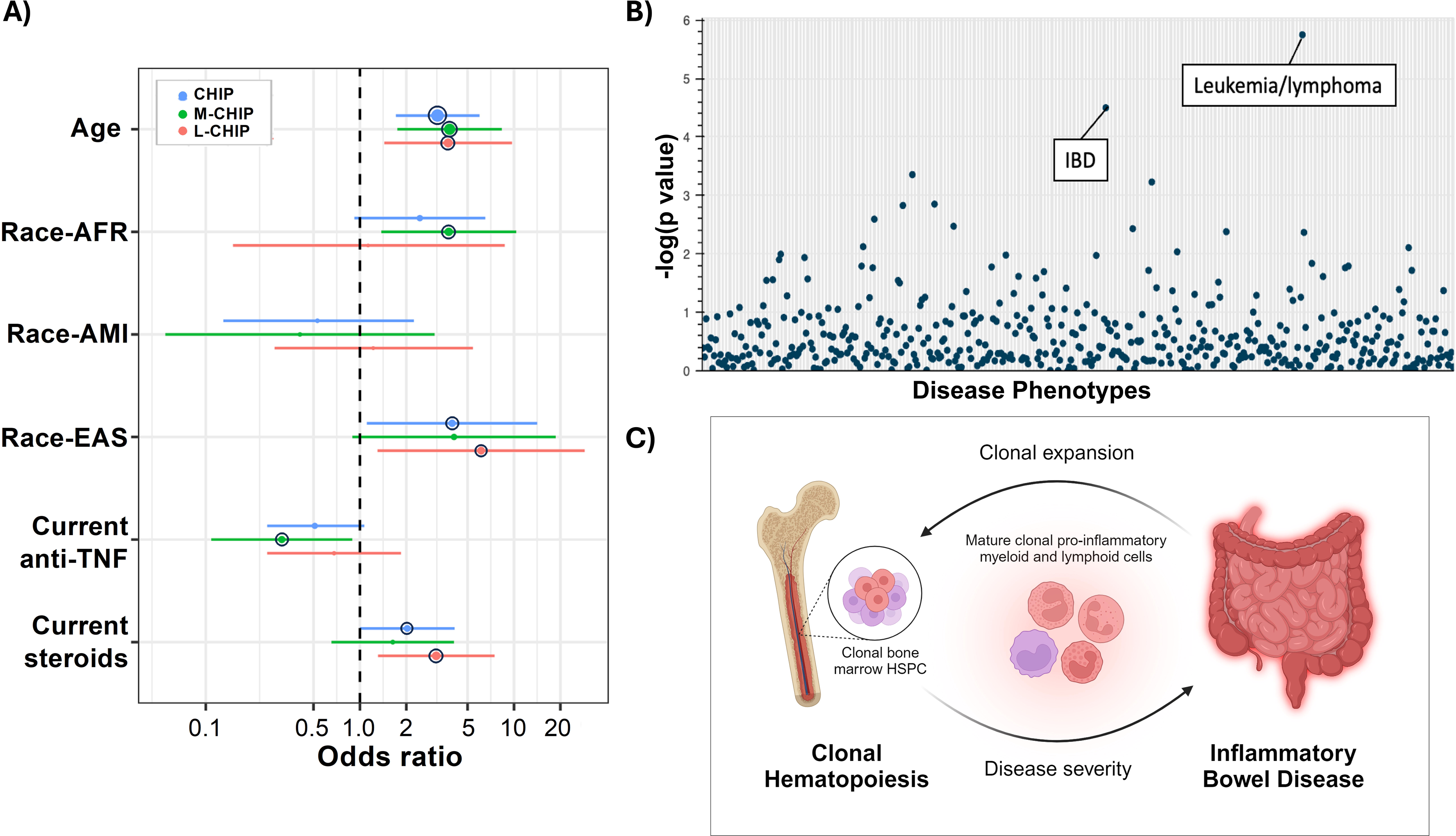
Highlighting connection between CHIP and IBD phenotype. A) Forest plot showing statistically significant variables identified in univariate analyses for increased risk of identifying CHIP, M-CHIP, and L-CHIP. The whiskers span the 95% confidence interval for OR values. The black circle outline indicates significance at p ≤ 0.05. B) PhenGenI plot of CHIP associated genes and clinical phenotypes. Degree of statistical significance of association is reflected by - log(p value). C) Hypothesized interplay of pro-inflammatory myeloid and lymphoid cell populations and inflammatory phenotype of IBD in which inflammation from mature CHIP-mutant immune cells propagate more severe disease phenotype and concurrent expansion of CHIP-mutant bone marrow hematopoietic stem and progenitor cells (HSPC). Figure created with Biorender.com. Alt text: A three paneled figure including a Forest plot of significant variables from a multivariate analysis, a dot-plot of phenotypes significantly associated with CHIP genes, and a hypothesized interplay between CHIP and IBD.

We did not observe a statistically significant association between CHIP and either current or past use of mercaptopurine (6-MP), azathioprine, or methotrexate which have been associated with the development of lymphoid/myeloid malignancies (Supplementary Table S5)^15, 16^. The lack of association between CHIP and medications traditionally associated with lymphoid/myeloid malignancies, and instead a link between CHIP and medications associated with treatment refractory IBD (corticosteroids), suggests CHIP mutations may evolve independent of mechanisms associated with medication toxicity and instead depend on mechanisms related to inflammation.

### Genome and phenotype analysis links IBD and hematologic malignancy

To understand potential mechanistic links between CHIP mutations and IBD, we queried our CHIP gene dataset against a database of publicly available genome and phenotype associations (PhenGenI)^22^. This analysis confirmed that the strongest association between CHIP genes and human disease was with leukemia/lymphoma, though the second most statistically significant associated disease was with IBD (Figure 6B). A similar analysis querying the CHIP gene dataset against a database of functionally annotated cellular pathways (KEGG) again identified a strong enrichment in pathways associated with cancer but also pathways associated with IBD pathophysiology and treatment including JAK-STAT and pI3K-AKT signaling (Supplementary Figure S1)^23, 24^. The substantial overlap in CHIP genes with those associated with the development and pathophysiology of IBD further reinforces that CHIP mutations may explain previously unknown associations between IBD and malignancy. Additionally, CHIP mutations may shape IBD outcomes related to disease onset and therapeutic response.

## Discussion

Here we present the results of a large single-center study of CHIP prevalence and characteristics in IBD patients and the first to include internal healthy controls and patients with CD. Our results reinforce previous findings in UC revealing a mutational spectrum that includes *DNMT3A, TET2, ASXL1, and PPM1D* variants dominating M-CHIP in UC. The inclusion of healthy controls in our cohort revealed a statistically significant increased prevalence of M-CHIP in older UC patients compared to healthy controls. The finding of an increased prevalence of M-CHIP mutations in older UC patients parallels the age distribution of CHIP in older patients with other inflammatory diseases including gout, vasculitis, and arthritis^7, 8, 17, 25^. These findings suggest that other factors specific to these inflammatory diseases may increase the development or CHIP or the perpetuation of CHIP clones. Older patients with UC may therefore be considered at higher risk for M-CHIP and associated malignant and non-malignant adverse outcomes including cardiovascular disease, stroke, and all cause mortality^3, 4, 26, 27^. Conflicting data exist regarding the specific risks of developing cardiovascular events in IBD as well as myeloid and lymphoid malignancies in IBD patients^28^. It may be that the presence of CHIP is a critical modifier that warrants evaluation in patients with UC that develop these complications.

Of particular clinical concern is our observation of high VAF M-CHIP in young IBD patients. The finding of high VAF M-CHIP in young IBD patients is notable as population level surveys, including over 500,000 individuals, demonstrate that CHIP is rare in individuals younger than 40 years of age^3, 4, 20^. The high clonal burden (i.e. high VAF) observed in young CD patients is the greatest risk factor for the eventual development of hematologic malignancy, as well as non-hematologic morbidity such as cardiovascular disease^2, 7, 10, 11, 27, 29^. Among the high-VAF clones, mutations in *TET2* were most common. Variants in *TET2* have been implicated in various models in upregulated inflammatory responses and competitive growth advantages of hematopoietic clones leading to cancer^6, 30–32^. Therefore, the detection of CHIP in younger IBD patients, and especially high-VAF *TET2* mutations, further raises concern for an increased risk of hematologic malignancy or cardiovascular complications over their lifetime. We did not observe incident cancers in the population of individuals with IBD and CHIP over a median follow up of 5.8 years. This interval, however, is likely too short to capture any expected progression which approaches 5% of all CHIP carriers over 10 years in retrospective observational cohorts^10, 11^.

Phenotypic and genomic analyses of CHIP genes demonstrated that the most highly associated phenotype after hematologic malignancy was IBD. The function of genes associated with CHIP are involved in various host-microbe interactions and inflammatory pathways, reinforcing that the development and pathophysiology of these two diseases may be intertwined. Systemic inflammatory consequences of CHIP are thought to arise from dysregulated circulating innate immune cells capable of reinforcing an inflammatory niche due to upregulation of cytokine production and a survival advantage of pro-inflammatory progenitor cells imparted by the CHIP mutations^6, 7, 30, 31, 33^. As such, there may be a bi-directional relationship between CHIP and IBD wherein each disease may reinforce the pathophysiology of the other (Figure 6C). In murine colitis models hematopoietic *Tet2* and *Dnmt3a* knock-out increased the severity of colitis and, in *Dnmt3a* knock-out, the development of colorectal cancer^32, 34, 35^. The potential that CHIP mutations may affect IBD clinical outcomes is suggested in our study where current steroid use was associated with an increased risk of CHIP whereas current anti-TNF therapy was associated with a decreased risk for CHIP. In the multivariate model steroid use remained a statistically significant risk-factor for the presence of CHIP suggesting an association between CHIP and patients with more severe steroid-dependent clinical phenotypes^36^. Understanding the interplay between IBD, CHIP and clinical IBD outcomes including the risk for severe disease will require validation in larger longitudinal cohorts. It will also be interesting to evaluate the mechanism by which medications used in IBD affect hematopoietic maturation and how this reinforces our observations linking CHIP and IBD^37, 38^.

Our study was limited in its retrospective nature and future prospective studies will be important to associate the presence of CHIP in IBD patients with clinical variables of interest. Our study cohort was further limited in lacking appropriate age-matched populations among CD, UC and control participants (median ages were 38 vs 44 vs 54 years, respectively) due to control participants being recruited for age-indicated procedures. The age differences among our populations and overall small numbers of CHIP carriers limited our ability to statistically assess important differences such as the prevalence of CHIP in young patients. Similarly, since the overall median age in our cohort was 20 years younger than previously published data of CHIP in UC, it likely resulted in a lower overall prevalence of CHIP in our study (4.6% vs 12.8%) given the expected increase of CHIP with age^3, 4, 17^. However, we caution against direct comparisons between studies given differences in sequencing platforms and lack of standards in filtering criteria and sequence coverage. Additionally, while there are no standard definitions of CHIP genes, we applied variants published in other database studies to our cohort analyses. Despite the younger age and lower overall prevalence seen in our study, we were still able to identify high risk cohorts among older UC and younger CD patients.

In summary, our study revealed an elevated risk for CHIP in older patients with UC, and younger CD patients with high clonal burdens potentially placing them at risk for CHIP associated complications. In multivariate phenotype analyses, surrogates for disease severity or control were aligned with the presence or absence of CHIP, respectively, suggesting a potential pathophysiological link. Our findings prompt clinically relevant hypotheses about the relationship between somatic alterations in hematopoietic cells, inflammation, treatment response and the risk for malignancy in IBD. In this context, CHIP may represent a novel variable to provide insights into known complications of IBD. These findings underscore the importance of awareness of CHIP among gastroenterologists and other clinicians. They provide rationale for the prompt referral of IBD patients with unexplained cytopenias to hematologists for comprehensive evaluation including potential bone marrow biopsy and genetic analysis.

Prospective longitudinal analyses are necessary to understand the consequences of CHIP in IBD patients and functional analyses will be crucial to elucidate the mechanistic interplay between hematopoietic somatic variants and the pathophysiology of IBD.

## Acknowledgements

We are grateful to Jill K Gregory, MFA, CMI for her assistance with designing and illustrating the graphical abstract.

## Supplementary Data

<u>Supplementary Table S1.</u> List of myeloid associated (M-CHIP) mutations and lymphoid associated (L-CHIP) mutations considered in this study.

<u>Supplementary Table S2.</u> List of M/L-CHIP mutations identified among the 587 CD patients, 441 UC patients and 293 non-IBD controls.

<u>Supplementary Table S3.</u> CHIP and age associations.

<u>Supplementary Table S4.</u> CHIP and VAF associations.

<u>Supplementary Table S5.</u> CHIP and demographics/disease characteristics associations in IBD patients.

<u>Supplementary Figure 1.</u> KEGG pathways are organized into colored clusters based on gene similarity. Gene enrichment analysis of the CHIP dataset identifies associations with KEGG pathways implicated in cancer, inflammation and host-microbe interactions supporting an overlap between IBD and cancer pathophysiology. Significant pathways (black circle) are labeled by name (corrected p<0.05).

