## Supplementary figures and images for "Clonal Hematopoiesis of Indeterminate Potential in Crohn’s Disease and Ulcerative Colitis"

### Supplementary Figure 1

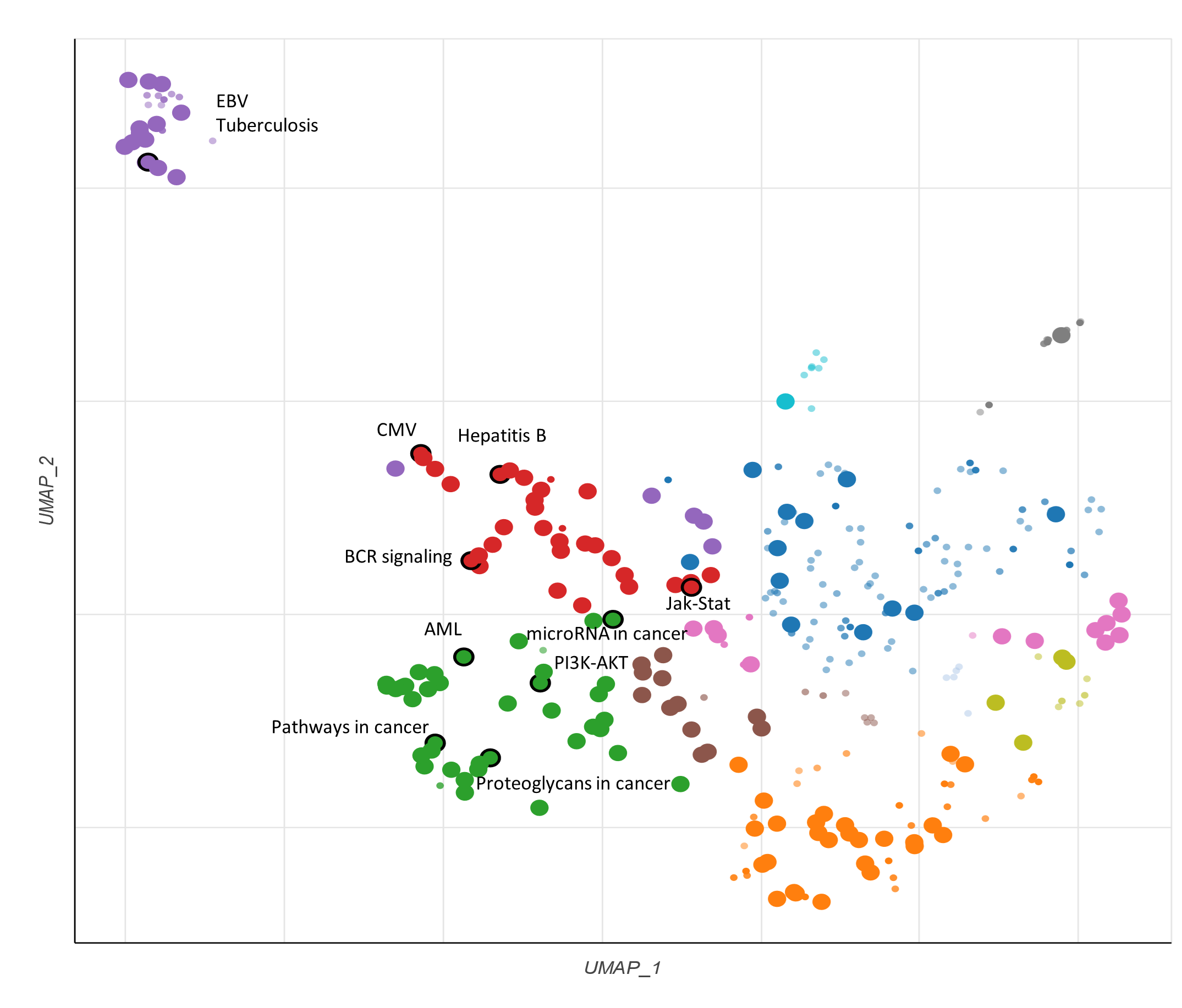
